# A pre-registered prospective study of large language models predicting late-breaking cardiovascular trial results at ESC Congress 2026

**DOI:** 10.64898/2026.09.23.26363773

**Authors:** Ki-Hyun Jeon, Ju-Seung Kwun, Hyoung-Won Cho

**Affiliations:** Division of Cardiology, Department of Internal Medicine, Seoul National University Bundang Hospital, Seongnam, Republic of Korea

**Keywords:** large language models, clinical trials, prediction, calibration, pre-registration, artificial intelligence

## Abstract

**Aims:** It is unknown whether large language models (LLMs) can predict a trial’s result at the design stage, from the information available when it is registered. We tested three LLMs prospectively on the Late-Breaking Science programme of the European Society of Cardiology (ESC) Congress 2026.

**Methods and results:** Of 139 trials, 50 were selected by pre-specified criteria, and predictions were locked and registered on the Open Science Framework before the congress. Claude Fable 5, GPT-5.5 and Gemini 3.5 Flash, without web access, received each title and an investigator-compiled summary of the registered design. They gave the probability that the primary endpoint would be met, and a point estimate with an 80% prediction interval for a locked effect measure. Results were adjudicated from publications (19) or presenters’ slides (31). Of 48 scorable trials, 28 (58%) were positive. The area under the receiver operating characteristic curve was 0.80 (95% CI 0.66–0.92) for Claude, 0.79 (0.66–0.91) for GPT and 0.74 (0.59–0.87) for Gemini, with accuracies of 73%, 71% and 67%. Among 26 trials whose effect was reported exactly as locked, the direction was correct in 88% of predictions and the median multiplicative error of ratio estimates was about 15%. The 80% prediction intervals contained the observed effect in 73.1% (Claude), 84.6% (GPT) and 57.7% (Gemini).

**Conclusion:** From the registered design alone, LLMs showed statistically significant discrimination between positive and negative trials and usually predicted the direction and approximate size of the effect. Such predictions might help trial design, and further studies are needed.

## Introduction

It is essential to predict the result when designing a clinical trial. That prediction informs the sample size and the choice of endpoint and comparator. Such predictions are not easy to make.[1,2] Large language models (LLMs) are increasingly studied for cardiovascular applications,[3] and ensembles of LLMs have matched human crowds on general prediction questions,[4] but we found no prospective test of their ability to predict clinical-trial results. Retrospective tests cannot exclude recall, because a model evaluated on trials reported before its training cut-off may have seen the answer.[5]

Late-breaking trials allow this to be tested prospectively. Their designs are registered publicly, and the results are embargoed until the day of presentation. We pre-registered predictions from three LLMs for the Late-Breaking Science programme of the European Society of Cardiology (ESC) Congress 2026 and scored them against adjudicated results.

## Methods

The protocol, design summaries, locked predictions and adjudication rules were registered on the Open Science Framework (OSF) before the congress (22 August 2026).[6] All 139 Late-Breaking Science trials were screened. Trials were excluded if the primary result was not a binary “met or not met” question (n=78), the results were already public (n=7), the registration or primary endpoint was unverifiable (n=1), or the title asserted the result (n=1). More trials were eligible (n=52) than the target of 50, so they were ordered by a rule fixed beforehand (randomized trials before secondary analyses of them, then trials with no related prior publication, then programme number) and the first 50 were taken, the two left out being secondary analyses. Thirteen of the 50 were new analyses of a published parent trial. For each trial the investigators compiled, before the congress, a structured design summary from the public registration of the presented trial or, for secondary analyses, its parent. Sources were ClinicalTrials.gov (45) and another national registry (3). Two pooled analyses had no registration, so their summaries used the component trials’ registrations and public design descriptions. Results-related information was excluded.

Claude Fable 5 (Anthropic, San Francisco, CA, USA), GPT-5.5 version 2026-04-23 (OpenAI, San Francisco, CA, USA) and Gemini 3.5 Flash (Google DeepMind, London, UK) were queried through their application programming interfaces (APIs). They received the title, the unchanged design summary, and investigator-written definitions of a positive result and the effect measure. They had no web access or tools, so all three worked from identical information and could not retrieve results. Predictions were generated on 21–22 August 2026 with default sampling settings. Each gave the probability (0–100) that the primary endpoint would be met, and a point estimate with an 80% prediction interval for the locked effect measure (e.g. hazard ratio, difference in proportions), answering three times in separate calls, and the median of the three answers was scored. When a model declined to answer all three times, the registered rule assigned it the uninformative midpoint of 50. The mean of the three models was defined as a fourth predictor before any result was adjudicated.

The investigator adjudicated results from the simultaneous publication (19 trials) or the presenters’ slides (31), using registered rules. Only numbers count, not the presenters’ wording, because spin is common in abstracts of cardiovascular trials with non-significant primary endpoints.[7] Non-inferiority trials are positive only if the pre-specified margin is met. The locked effect measure is scored only when reported as locked. A trial whose locked endpoint is not presented is not scored.

The registered primary outcomes were discrimination of the probabilities and, for the quantitative predictions, interval coverage and effect-size error. Discrimination is reported as the area under the receiver operating characteristic curve (AUC), with calibration, accuracy, sensitivity and specificity at a threshold of 50.[8] Ratio predictions given on a log scale were exponentiated by a registered rule. Confidence intervals and P values are from a bootstrap (4000 resamples). Proportions use Wilson intervals, and no adjustment was made for multiplicity. A majority vote, a wider quantitative set and a correction of predictions given on another scale were added post hoc. Analyses used Python 3.12 (Python Software Foundation, Wilmington, DE, USA), and reporting follows TRIPOD-LLM where applicable.[9]

## Results

Two trials could not be scored, because a different trial was presented in one slot and the locked dose was not reported in the other. Of the other 48, 28 (58%) were positive.

The AUC (Figure 1A) was 0.80 (95% CI 0.66–0.92) for Claude, 0.79 (0.66–0.91) for GPT, 0.74 (0.59–0.87) for Gemini and 0.78 (0.64–0.91) for the ensemble, each above chance (P<0.001, P<0.001, P=0.004 and P<0.001). Claude discriminated better than Gemini (difference in AUC 0.065, 95% CI 0.003 to 0.139, P=0.042), whereas the differences between Claude and GPT (0.007, ™0.041 to 0.059, P=0.83) and between GPT and Gemini (0.058, ™0.005 to 0.139, P=0.08) were not statistically significant. Accuracy was 73% (59–83) for Claude, 71% (57–82) for GPT and 67% (53–78) for Gemini. The post hoc majority vote reached 71% (57–82), with sensitivity 22/28 (79%) and specificity 12/20 (60%). Calibration slopes were 0.84, 1.18 and 0.55, all with confidence intervals including 1. The same models had also predicted from the title alone, before the design summary, with lower AUCs (0.70, 0.67 and 0.65) and accuracy of 67%, 63% and 65%.

**Figure 1.**
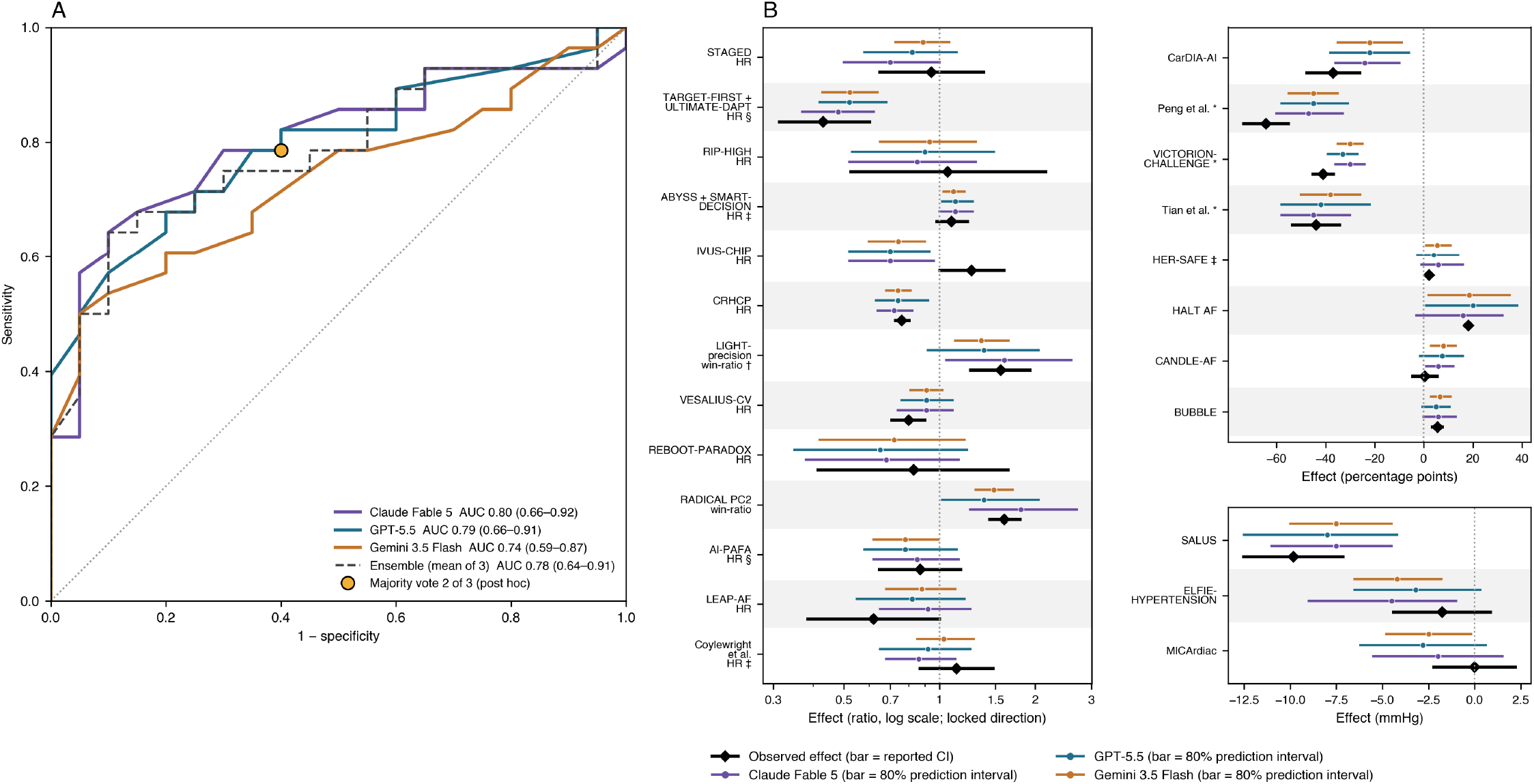
Predictions of ESC 2026 late-breaking trial results by three large language models.s. **(A)** ROC curves for the predicted probability that the primary endpoint would be met. **(B)** Predicted effect sizes with their 80% prediction intervals, compared with the observed effects, for 24 of the 26 trials whose unit is shared by at least three trials. The effect measure is given beneath the ratio estimands. Observed confidence intervals are 95% unless marked. None is shown for HER-SAFE, which reported only the upper limit of a 90% interval, or for HALT AF, which reported an unadjusted difference and no interval. Predictions are drawn after a post hoc scale correction, whereas the coverage quoted in the text uses the pre-specified aggregation. For two trials the locked measure differs from the primary endpoint reported. For TARGET-FIRST and ULTIMATE-DAPT pooled it was clinically relevant bleeding (BARC 2, 3 or 5) rather than BARC 3 or 5 bleeding, and for VESALIUS-CV all-cause death rather than the dual primary composite endpoints. † 97.5% confidence interval. ‡ Registered as a non-inferiority question, so success required a pre-specified margin rather than the null line. §Presented as a non-inferiority study but scored against the locked superiority question. *Percent change in a biomarker. ROC, receiver operating characteristic; BARC, Bleeding Academic Research Consortium.

Agreement between models (κ 0.69–0.82) exceeded their agreement with the adjudicated results (κ 0.30–0.44). Of 26 trials that all three called positive, 21 were positive, and of 13 that all three called negative, 8 were negative. Claude declined to answer for one trial.

The effect measure was reported exactly as locked for 26 trials (Figure 1B). With the pre-specified aggregation, the direction of effect was correct in 22/25 (88%) predictions for each model. For the 13 trials whose effect was a ratio, such as a hazard ratio, half of the predictions were within about 15% of the observed value. The 80% prediction intervals contained the observed effect in 19/26 (73.1%, 95% CI 53.9–86.3) for Claude, 22/26 (84.6%, 66.5–93.8) for GPT and 15/26 (57.7%, 38.9–74.5) for Gemini. An exploratory analysis of 40 trials, also accepting effects computed from the arm-level numbers presented (8) or reported with a different estimator (6), gave coverage of 74.4%, 80.0% and 52.5%.

Several sensitivity analyses were run, scoring the mis-mapped trial on what was actually presented, excluding the refused trial, and analysing separately the trials with and without a published parent and those adjudicated from publications or slides. Gemini ranked last in all of them, but the order of Claude and GPT was not stable. Excluding the six trials whose presented endpoint or timepoint differed from the one locked, as the registered rule required, left 42 trials with AUCs of 0.79, 0.80 and 0.74.

## Discussion

This study investigated whether LLMs can predict the result of a cardiovascular trial from its registered design. Three frontier models received the title and design summary of each late-breaking trial at a major congress, and their predictions were locked before the congress opened. All three showed statistically significant discrimination between positive and negative trials, and they usually predicted the direction and approximate size of the treatment effect. The models tended to be wrong on the same trials, so agreement between them is not independent confirmation, and the 80% intervals did not reach 80% coverage in every model.

Oncologists predicting trials in their own subspecialty could not separate positive from negative trials, individually or in aggregate (AUC 0.52 and 0.43),[1] and principal investigators predicting their own trials were over-optimistic although they discriminated modestly (AUC 0.76).[2] The AUCs of 0.74 to 0.80 here are therefore at least as high as either group achieved. Earlier tests of LLMs on clinical trials have been retrospective. Windisch et al. found that models identified the outcome of published oncology trials from the title alone with 79% to 88% accuracy, and beat guessing the commoner outcome when given only the article identifier.[5] In the present study every prediction was locked and registered before any result was public, so recall cannot have contributed. The models also explained their reasoning, each comparing the registered design with published trials of similar interventions and estimating the effect size from them. Half of the supporting claims they made were also made by another model.

This study has limitations. A single investigator, who had seen the predictions, adjudicated the results under registered rules. Two trials had been matched to the wrong registry record. Six were scored although the presented endpoint or timepoint differed from the one locked and excluding them did not change the conclusions. Only half of the trials reported the effect measure exactly as locked.

In conclusion, a prospective pre-registered test showed that LLMs can predict the results of late-breaking cardiovascular trials from the design information available at registration, although they are not yet reliable enough to stand in for an expert’s own prediction. A dedicated test is needed to show whether such predictions help trial design, and whether the finding holds for later models.

## Data Availability

The pre-congress registration (protocol as registered, design summaries, 1,200 locked prediction files) is available on the Open Science Framework at https://osf.io/w6328 (doi 10.17605/OSF.IO/W6328). The post-congress component (protocol with the dated post-registration record, the locked adjudication file with SHA-256 checksums, and the scoring outputs for all registered measures) is available at https://osf.io/346xn.

## Data availability

The pre-congress registration (protocol as registered, design summaries, 1,200 locked prediction files) is at https://osf.io/w6328 (doi 10.17605/OSF.IO/W6328). The post-congress component is at https://osf.io/346xn (uploaded 18 September 2026). It holds the protocol with the dated post-registration record, the locked adjudication file with the SHA-256 checksums recorded when it was locked, and the scoring outputs for all registered measures.

## Ethics

This study used only publicly available, trial-level information (congress programme, trial registry records, publications and presentation slides) and the outputs of language models. No human participants, patient-level data or animals were involved. Ethics committee approval and informed consent were therefore not required.

## Consent

Not applicable. The study involved no human participants or patient-level data, so informed consent was not required.

## Author contributions

K.-H.J. conceived and designed the study, generated and locked the predictions, adjudicated the results, performed the analysis, and drafted the manuscript. J.-S.K. and H.-W.C. contributed to the interpretation of the data and critically revised the manuscript. All authors approved the final version and agree to be accountable for the work.

## Use of artificial intelligence

The three language models evaluated are the subject of the study. In conducting and reporting it, the authors also used Claude Opus 5 and Claude Fable 5.1 (Anthropic, through Claude Code) and GPT-6 Astra (OpenAI, through Codex). All outputs were checked by the authors, who take full responsibility for the content.

